# No change in 24-hour salt intake estimated from spot urine in Norwegian adults from 2006 to 2019. The population-based HUNT Study

**DOI:** 10.1101/2023.08.14.23294065

**Authors:** Kristin Holvik, Marianne Hope Abel, Jostein Holmen, Steinar Krokstad, Torunn Holm Totland, Haakon E Meyer

## Abstract

**Objective:** Monitoring time trends in salt consumption is important for evaluating the impact of salt reduction initiatives on public health outcomes. There has so far not been available data to indicate if salt consumption in Norway has changed during the previous decade. We aimed to assess whether average 24-hour salt intake estimated from spot urine samples in the adult population of mid-Norway changed from 2006-08 to 2017-19, and to describe variations by sex, age, and educational level.

**Design:** Repeated cross-sectional studies.

**Setting:** The population-based HUNT Study.

**Participants:** In each of two consecutive waves (HUNT3: 2006-08 and HUNT4: 2017-19), spot urine samples were collected from 500 men and women aged 25-64 years, in addition to 250 men and women aged 70-79 years in HUNT4. Based on spot urine concentrations of sodium, potassium and creatinine, age, sex, and body mass index, we estimated 24-hour sodium intake using the INTERSALT equation for the Northern European region.

**Results:** Mean (95% confidence interval (CI)) estimated 24-hour salt intakes in men were 11.1 (95% CI 10.8, 11.3) g in HUNT3 and 10.9 (95% CI 10.6, 11.1) g in HUNT4, p=0.25.

Corresponding values in women were 7.7 (95% CI 7.5, 7.9) g and 7.7 (95% CI 7.5, 7.9) g, p=0.88. Mean estimated salt intake in HUNT4 decreased with increasing age in women, but not in men, and it did not differ significantly across educational level in either sex.

**Conclusions:** Estimated 24-hour salt intake in adult men and women in mid-Norway did not change from 2006-08 to 2017-19.

## Introduction

Excess sodium consumption is linked to hypertension and risk of cardiovascular disease (CVD), and a shift towards a lower sodium to potassium ratio (Na/K ratio) in the diet has shown improved outcomes in terms of lowering blood pressure and reducing cardiovascular morbidity and mortality ^(1-6)^. A global initiative to reduce dietary salt intake for improving cardiovascular health is coordinated by the World Health Organization (WHO) ^(7)^. In all member states of the WHO European Region where sodium intakes have been assessed by urine collections, average intakes clearly exceed the recommended maximum level of 2 g/day for adults, corresponding to approximately 5 g salt ^(8-10)^. Several countries, including Norway, have committed to reduce population salt intakes by 30% between 2010 and 2025 ^(7, 11)^. Strategies to achieve this goal involve reductions of salt content in processed foods and increasing consumers’ awareness through information and food labelling. National policies that involve structural interventions have a potential for reducing population salt intake ^(12)^. The interventions should be implemented gradually to allow for adapting to the less salty taste of foods, thus preventing consumers from shifting to saltier products or compensating by adding more salt during preparation and consumption. As in several countries, there has been progress in Norway ^(13)^ with reduced salt content in common foods through a commitment between food industry and the government ^(14)^. Monitoring time trends in salt consumption is necessary for evaluating the public health impact of these initiatives. In the large population-based HUNT Study in mid-Norway, systolic blood pressure declined sharply from the mid-1990s to the mid-2000s ^(15)^ and thereafter there was a continued slight decline until 2017-2019 ^(16, 17)^. There has so far not been available data to indicate if population salt consumption has changed during the last decade.

Approximately 90% of ingested sodium is excreted through the urine, and measurements of 24-hour urinary sodium excretion in representative samples is recommended as an indicator of average population sodium intake ^(10, 18)^. However, collecting 24-hour urine is resource-demanding and collection of spot urine samples may often be more feasible. Therefore, equations for predicting 24-hour excretion from spot urine concentrations have been developed in different populations ^(19-22)^. A meta-analysis of validation studies concluded that the mean population 24-hour salt intake can be predicted fairly accurately from spot urine samples collected in the same population ^(23)^. In a population-based study in Northern Norway, excretion of sodium, potassium and creatinine was measured in both 24-hour urine and spot urine ^(24)^. Among three previously developed equations for estimating salt intake from spot urine concentrations, the equation developed in the International Cooperative Study on Salt and Blood Pressure (INTERSALT) study with the coefficient for the Northern European region ^(20, 25)^ yielded the most accurate estimate of average 24-hour sodium excretion and thus represents the preferred choice in an adult Norwegian population ^(24)^.

Urinary sodium, potassium, and creatinine concentrations were available from spot urine samples of participants in two waves of a large population-based health study in mid-Norway in 2006-08 (n=500 aged 25-64 years) and 2017-19 (n=500 aged 25-64 and n=250 aged 70-79 years). Participant sampling, urine collection and biochemical analyses were performed using identical methods in both waves ^(17)^. We aimed to assess whether there has been a change in salt intake during the period, and to assess possible variations in urinary concentrations of sodium, potassium, Na/K ratio, and estimated salt intake across sex, age, and educational level.

## Methods

### Study population

The HUNT Study is a population-based cohort study of the adult population in Trøndelag County in mid-Norway, a region characterized by a relatively homogenous ethnic population and low migration, suitable for examining time trends ^(17)^. The study has been running in Nord-Trøndelag since 1984, inviting all inhabitants aged 20 years and older to participate. The data collection covers a wide range of health-related topics through repeated surveys with questionnaires, interviews, clinical examinations, laboratory measurements and stored biological samples. The third wave (HUNT3) was carried out in 2006-08 and the fourth wave (HUNT4) was carried out in 2017-19. Overall participation rate, defined as completing the main questionnaire and attending the clinical screening ^(26)^, was the same in these two waves; 54% (49% in men and 59% in women). Spot urine samples were collected from approximately 12000 participants in HUNT3 and 26900 participants in HUNT4. For our purpose, age- and sex-stratified subsets of 500 HUNT3 participants and 750 HUNT4 participants were drawn randomly from participants who had donated a spot urine sample and had no known kidney disease, heart failure or cerebral stroke. These comprised 125 individuals of each sex in each of the age strata 25-44 years and 45-64 years in both waves, and an additional 125 individuals of each sex aged 70-79 years who attended the HUNT 70+ extension of HUNT4.

### Biochemical analyses

In both waves, spot urine samples were non-fasting and taken any time during the day (9am to 8pm). The samples were immediately cooled down to 4°C, transported at 4°C to HUNT Biobank the same day and frozen to -80°C the following day for storage. The analyses were performed at Levanger Hospital with Architect ci8200. Urinary sodium and potassium were assessed with an indirect ion-selective electrode to determine ion concentration. Urinary creatinine was assessed using an enzymatic assay. Measurement range was 220 - 35360 μmol/L. Total assay Coefficients of Variability (CV) was 24% (5790 μmol/L).

### Covariates

Height was measured to the nearest centimetre and weight was measured to the nearest half-kilogram without shoes and wearing light clothing. Body mass index (BMI) was calculated as body weight in kilograms divided by the squared value of body height in meters (kg/m^2^). In HUNT4, self-reported educational attainment was collected through a questionnaire asking for the highest completed education, with six response options ranging from primary school to college/university education of ≥4 years, based on the Norwegian Standard Classification of Education ^(27)^. Response options 1 and 2 and response options 3 and 4 were combined, resulting in four categories: Primary education (completed primary school or 1-2 years of high school/vocational school), secondary education (completed high school or certificate of apprenticeship), university or other post-secondary education <4 years, and university/college education of ≥4 years. For studying differences by age, individual age was categorized into 25-44 years, 45-64 years, and 70-79 years.

### 24-hour salt intake

Urinary concentrations of sodium, potassium, and creatinine were determined in mmol/l. Na/K ratio was defined as urinary sodium concentration (mmol/l) divided by urinary potassium concentration (mmol/l). The INTERSALT equation with coefficients for the Northern European region ^(20)^ was used to estimate 24-hour sodium excretion. This equation has been validated and found to perform well in the age range 40-69 years in a population-based study carried out in a geographically close Norwegian city with a comparable population to that in HUNT in terms of a homogenous ethnic background ^(24)^. The equation is shown in Supplementary Figure 1. It is separate for men and women and includes age (years), BMI (kg/m^2^), and spot urine concentrations of sodium, potassium and creatinine (mmol/l). To convert from mmol to g Na/24h, results were multiplied by a factor of 0.023, then multiplied by a factor of 2.54 to obtain NaCl (g/24h).

### Statistical analysis

Statistical analysis was performed in Stata SE version 17.0 (Stata Corp., College Station, TX). All analyses were performed in the age groups represented in both waves (25-44 and 45-64 years). Participants aged 70-79 years were included only when describing mean values by age in HUNT4. Differences in mean values between HUNT3 and HUNT4 were tested with two-sided T-test. We performed linear regression and used the *margins* command to estimate mean values with standard deviations and 95% confidence intervals (CI) within groups of sex, age, and education level. Mean estimated salt intakes (g/24h) by educational level were age-standardized by including individual age as a covariate in the regression model and stratified by sex. Differences between age groups were tested by analysis of variance (ANOVA), while differences between groups of educational level were tested by analysis of covariance (ANCOVA) taking age into account. For results presented as supplementary material, median values by sex, age, and educational level were obtained by performing quantile regression followed by the *margins* command in Stata.

## Results

### Characteristics of the study population

We included 499 HUNT3 participants and 500 HUNT4 participants aged 25-64 years, and an additional 250 participants aged 70-79 years in HUNT4. Mean characteristics in the age span represented in both waves (25-64 years) are shown in Table 1. The corresponding median values are shown in Supplementary Table 1. Mean age was 46 years in both waves as a result of the age-stratified sampling (Table 1). BMI in men 25-64 years old was similar in HUNT4 as in HUNT3: mean 27.6 vs. 27. 8 kg/m^2^, p=0.55. In women 25-64 years old, BMI tended to be slightly higher in HUNT4, mean 26.7 vs. 25.9 kg/m^2^ in HUNT3, p=0.09.

**Table 1.** Characteristics of men and women aged 25-64 years with spot urine samples in the HUNT Study: HUNT3 (2006-08, n=499) and the HUNT4 (2017-19, n=500). Mean (standard deviation)

|  | Men |  | P value† | Women |  | P value† |
| --- | --- | --- | --- | --- | --- | --- |
|  | HUNT3 | HUNT4 |  | HUNT3 | HUNT4 |  |
| Age | 46.8 (10.9) | 45.8 (12.0) | 0.33 | 46.1 (10.8) | 45.6 (11.9) | 0.63 |
| BMI | 27.8 (3.9) | 27.6 (4.4) | 0.55 | 25.9 (4.6) | 26.7 (5.2) | 0.086 |
| Na (mmol/l) | 122.1 (54.0) | 118.5 (52.5) | 0.45 | 94.7 (56.5) | 93.4 (57.0) | 0.80 |
| K (mmol/l) | 68.3 (31.4) | 74.3 (37.4) | 0.054 | 59.1 (32.3) | 62.7 (36.5) | 0.24 |
| Na/K ratio | 2.08 (1.18) | 1.92 (1.11) | 0.1813 | 1.83 (1.02) | 1.77 (1.21) | 0.53 |
| Creatinine | 12.0 (6.7) | 11.9 (6.3) | 0.87 | 8.9 (5.9) | 8.9 (6.5) | 0.87 |
| Calculated NaCl (g/24h)* | 11.05 (1.90) | 10.85 (1.86) | 0.25 | 7.69 (1.29) | 7.71 (1.35) | 0.88 |

BMI, body mass index. Na, urinary sodium concentration. K, urinary potassium concentration. Na/K ratio, ratio of urinary sodium to potassium concentrations. NaCl, salt.
\* Estimated using the INTERSALT equation with sex-specific coefficients for the Northern European region <sup>(20)</sup>
† Independent samples T test for mean difference between HUNT3 and HUNT4 within sex

### Sodium, potassium, and Na/K ratio in HUNT3 and HUNT4

Mean urinary sodium concentrations in adults aged 25-64 years were not significantly different in HUNT3 and HUNT4 (Table 1). In men, urinary potassium concentrations tended to be slightly higher in HUNT4 compared with HUNT3: mean 74 vs. 68 mmol/l (p=0.054). The difference in urinary potassium in women was small and not statistically significant, mean 63 vs. 59 mmol/l (p=0.24). Urinary Na/K ratio tended to be slightly lower in HUNT4 than in HUNT3 but the difference was not statistically significant: Mean 1.92 vs. 2.08 in men (p=0.13) and 1.77 vs. 1.83 in women (p=0.53) (Table 1). Median values showed a similar pattern (Supplementary Table 1).

### Estimated salt intake in HUNT3 and HUNT4

Among adults aged 25-64 years, mean estimated 24-hour salt intake in HUNT3 was 11.1 g (95% CI 10.8 g, 11.3 g) in men and 7.7 g (95% CI 7.5 g, 7.9 g) in women (Table 1). In HUNT4, the corresponding values were 10.9 (95% CI 10.6, 11.1) g and 7.7 (95% CI 7.5, 7.9) g, representing no change from HUNT3 to HUNT4. Results were similar for median values (Supplementary Table 1). In men and women combined (not shown in table), mean estimated salt intake was 9.37 g in HUNT3 and 9.28 g in HUNT4 (p=0.52), i.e., a difference of -0.09 g (95% CI -0.38 g, 0.20 g). Age and sex adjustment did not affect the result.

### Variations across sex, age, and educational level

Gradients across age and educational level are described for HUNT4 only. Overall and within categories of age and education, women had a lower estimated 24-hour salt intake than men. In women there was a clear age gradient, with mean estimated salt intake being highest in younger adults (8.1 g) and lowest in the 70-79-year-old women (5.9 g, p<0.001) (Table 2). In men, estimated 24-hour salt intake was similar across age (mean 10.7 g for ages 25-44, 11.0 g for ages 40-64, and 11.1 g for ages 70-79, p=0.24 in ANOVA). While urinary potassium concentrations were similar across age groups (Table 2), there was an age gradient in urinary sodium concentrations in both men and women, with the highest concentrations in the younger adults. Consequently, urinary Na/K ratio was higher in those aged 25-44 years, with a ratio above 2 in both men and women. In age groups ≥45 years, Na/K ratio was approximately 1.7 in men and below 1.6 in women (Table 2). In participants aged 25-64 years, 23% had completed college/university education of at least 4 years, 26% had completed college/university education of less than 4 years, 36% had completed secondary school, and 15% reported primary school as their highest completed education. Age-standardized mean estimated 24-hour salt intake did not differ significantly across educational attainment (Table 3). The corresponding median values are shown in Supplementary Tables 2 and 3.

**Table 2.**
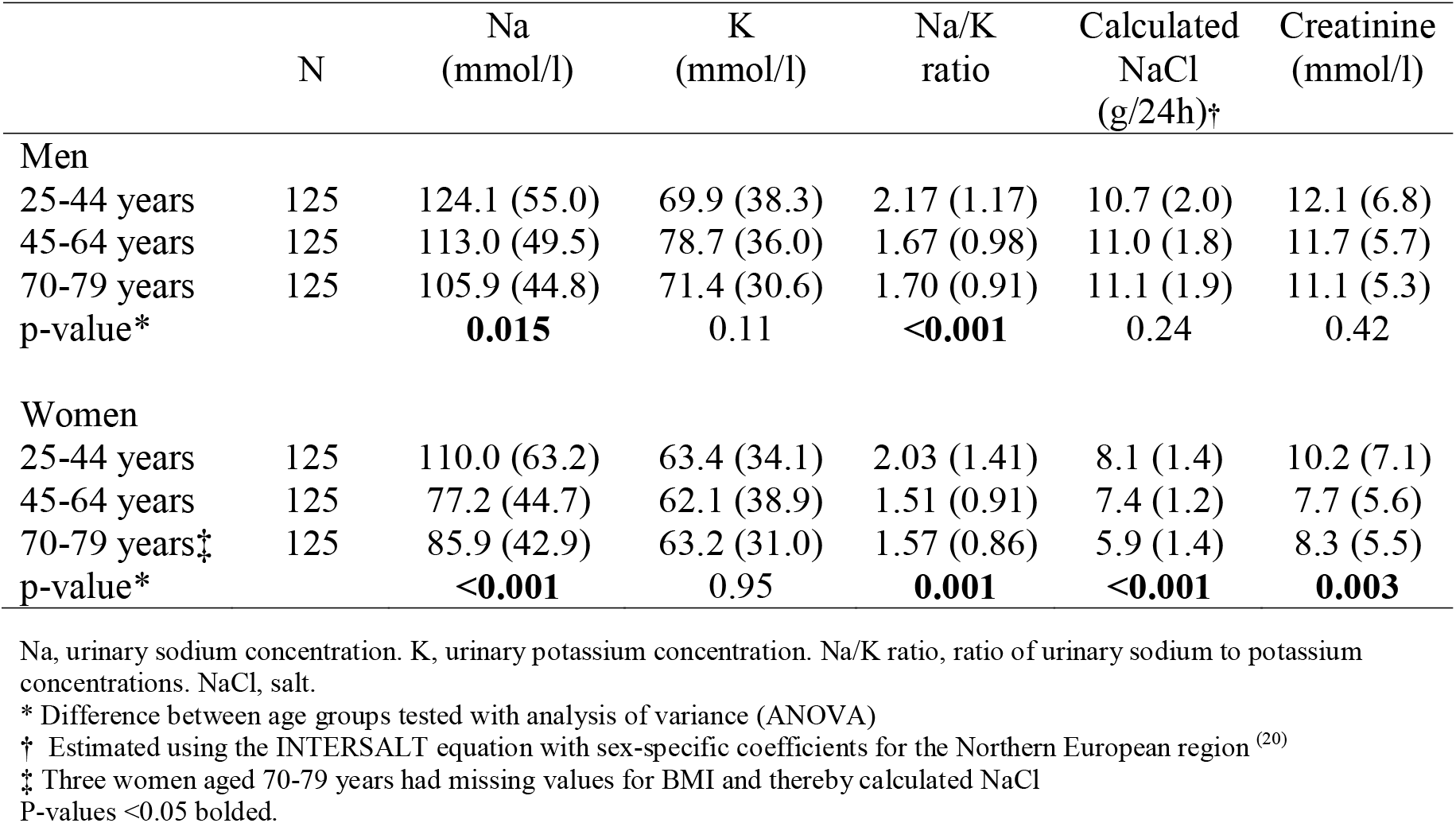
Spot urinary sodium (Na), potassium (K), sodium to potassium ratio (Na/K ratio), creatinine and estimated 24-hour salt intake by age groups in men and women participating in HUNT4 (n=750). Mean (standard deviation)

|  | N | Na (mmol/l) | K (mmol/l) | Na/K ratio | Calculated NaCl (g/24h)† | Creatinine (mmol/l) |
| --- | --- | --- | --- | --- | --- | --- |
| **Men** |  |  |  |  |  |  |
| 25-44 years | 125 | 124.1 (55.0) | 69.9 (38.3) | 2.17 (1.17) | 10.7 (2.0) | 12.1 (6.8) |
| 45-64 years | 125 | 113.0 (49.5) | 78.7 (36.0) | 1.67 (0.98) | 11.0 (1.8) | 11.7 (5.7) |
| 70-79 years | 125 | 105.9 (44.8) | 71.4 (30.6) | 1.70 (0.91) | 11.1 (1.9) | 11.1 (5.3) |
| p-value* |  | **0.015** | 0.11 | **<0.001** | 0.24 | 0.42 |
| **Women** |  |  |  |  |  |  |
| 25-44 years | 125 | 110.0 (63.2) | 63.4 (34.1) | 2.03 (1.41) | 8.1 (1.4) | 10.2 (7.1) |
| 45-64 years | 125 | 77.2 (44.7) | 62.1 (38.9) | 1.51 (0.91) | 7.4 (1.2) | 7.7 (5.6) |
| 70-79 years‡ | 125 | 85.9 (42.9) | 63.2 (31.0) | 1.57 (0.86) | 5.9 (1.4) | 8.3 (5.5) |
| p-value* |  | **<0.001** | 0.95 | **0.001** | **<0.001** | **0.003** |

Na, urinary sodium concentration. K, urinary potassium concentration. Na/K ratio, ratio of urinary sodium to potassium concentrations. NaCl, salt.
\* Difference between age groups tested with analysis of variance (ANOVA)
† Estimated using the INTERSALT equation with sex-specific coefficients for the Northern European region <sup>(20)</sup>
‡ Three women aged 70-79 years had missing values for BMI and thereby calculated NaCl
P-values <0.05 bolded.

**Table 3.** Age-standardized mean urinary sodium (Na), potassium (K), sodium to potassium ratio (Na/K ratio) and estimated 24-hour salt intake by educational attainment in men and women aged 25-64 years participating in HUNT4.

| Education level* | N | Na<br>(mmol/l) | K<br>(mmol/l) | Na/K<br>ratio | Calculated<br>NaCl<br>(g/24-hour) † |
| --- | --- | --- | --- | --- | --- |
| **Men** |  |  |  |  |  |
| Primary | 38 | 125.2 | 72.7 | 2.19 | 11.1 |
| Secondary | 110 | 122.4 | 75.7 | 2.04 | 10.9 |
| Tertiary, short (<4 yrs) | 59 | 115.6 | 79.9 | 1.66 | 10.6 |
| Tertiary, long (≥4 yrs) | 42 | 108.0 | 65.2 | 1.74 | 10.9 |
| p-value‡ |  | 0.38 | 0.26 | **0.045** | 0.52 |
| **Women** |  |  |  |  |  |
| Primary | 39 | 102.4 | 67.9 | 1.65 | 7.7 |
| Secondary | 67 | 98.8 | 59.4 | 2.00 | 8.0 |
| Tertiary, short (<4 yrs) | 72 | 82.2 | 63.4 | 1.59 | 7.5 |
| Tertiary, long (≥4 yrs) | 71 | 95.3 | 62.7 | 1.80 | 7.6 |
| p-value‡ |  | 0.19 | 0.73 | 0.20 | 0.23 |

Na, urinary sodium concentration. K, urinary potassium concentration. Na/K ratio, ratio of urinary sodium to potassium concentrations. NaCl, salt.
\* Attained educational level in four categories: Primary education: Completed primary school or 1-2 years of high school/vocational school; secondary education: Completed high school or certificate of apprenticeship; tertiary short: University or other post- secondary education less than 4 years; tertiary long: University/college education of 4 years or more.
† Estimated using the INTERSALT equation with sex-specific coefficients for the Northern European region <sup>(20)</sup>
‡ P-values obtained by linear regression followed by the *testparm* option.
P-values <0.05 bolded.

## Discussion

When estimating 24-hour salt intake from spot urine samples using the INTERSALT equation, we found that salt intake in adult men and women in mid-Norway did not change significantly from 2006-08 to 2017-19. We observed an overall difference in estimated salt intake of -0.09 g between the two time points, with 95% CI ranging from -0.38 to 0.20 g. The results suggest that salt consumption did not decrease in adults in mid-Norway during this period. There was a slight decline in blood pressure levels in this population during the same period ^(16, 17)^, suggesting that blood pressure may have been affected by changes in other risk factors. The magnitude of 24-hour salt intake was comparable to that in adults aged 40-69 years in the population-based Tromsø Study in 2015-16 (10.4 g in men and 7.6 g in women) ^(24)^.

### Estimating 24-hour salt intake from spot urine samples

For monitoring average sodium intake in populations, 24-hour urinary assessment is the recommended method ^(18)^. Dietary assessment methods have shown large measurement errors due to reporting errors, highly variable sodium content of food products which challenges food composition databases, and the difficulty to capture the amount of salt added during preparation and consumption ^(28-30)^. Spot urine samples are often available in large population-based studies, and equations to estimate 24-hour salt excretion have been developed ^(19-22)^.

In the population-based Tromsø Study 2015-16, both 24-hour urine excretion and spot urine concentrations of sodium, potassium and creatinine were measured in 475 men and women aged 40-69 years ^(24)^. Among three previously developed equations for estimating average 24-hour sodium intake from spot urine concentrations ^(19, 20, 22)^, the INTERSALT equation with the coefficients for the Northern European region yielded the most accurate estimate ^(20, 24)^. The mean 24-hour sodium excretion estimated by the INTERSALT equation was 4% higher in men and 1% lower in women than that measured by 24-hour urine, corresponding to an overestimation of 0.4 g salt in men and an underestimation of 0.1 g salt in women ^(24)^. The similarity of our findings in the HUNT population to those of the Tromsø population supports the generalizability of our results in adults aged <70 years. A previous validation study in a US population aged 18-39 years also found the INTERSALT equation to perform well among several equations examined ^(31)^, but its validity is not clarified in older adults ^(32)^. Furthermore, the INTERSALT equation tends to underestimate 24-hour NaCl at high intakes and overestimate NaCl at low intakes ^(20)^, with the inherent risk of failing to detect a true decline. However, reassuringly, we observed that sodium concentrations in spot urine were also unchanged and showed in general the same pattern across subgroups of sex, age, education, and study wave, supporting a true null finding.

### Variations by age

There was a clear age gradient in both men and women in 2017-19, with higher urinary sodium concentrations in the youngest participants. Estimated salt intake was lowest in the oldest women, while in men there was no significant difference by age. This is in line with findings from the Tromsø study where calculated salt intake based on 24-hour urine collection was inversely related to age in women but not in men ^(24)^. We may speculate that the finding of higher salt intakes in younger compared with older women may reflect a more frequent consumption of processed foods, take-away and readymade foods with a higher salt content in the younger age groups, at the expense of home-made food prepared from raw ingredients ^(33)^. Furthermore, salt intake may be lower in older women both due to a lower energy intake and a greater attention to sources of salt. Notably, a nationwide online public health survey conducted in people aged 18 years and older in Norway in 2020 ^(34)^ showed an age gradient in the proportion who reported to add additional salt “often or usually” to food during preparation and serving, with higher proportions in younger adults. While the proportion was up to 30 percent in the younger age groups (up to 54 years), it was lower in those 55 years and older, and lowest in women aged >75 years (11%). In addition, the proportion who reported frequent consumption of potato chips and salty snacks was highest in younger age groups (18-49 years). Younger age was a stronger predictor than geography and education for both salt added to meals and frequency of salty snacks consumption ^(34)^.

### Urinary Na/K ratio

Mean urinary Na/K ratio in people aged 25-64 years in 2017-19 was approximately 1.9 in men and 1.8 in women. Urinary Na/K ratio was highest (above 2) in younger adults. This difference reflected the age difference in sodium concentrations, while potassium concentrations did not exhibit a trend across age. Na/K ratio can be lowered both by lowering sodium through salt reduction and by increasing potassium, e.g., through increased fruit and vegetable consumption, and is proposed to be the most relevant measure in relation to blood pressure and CVD risk ^(35)^. While there is no officially recommended target value for Na/K ratio, meeting the dietary target intake of sodium would result in a ratio of approximately 1 ^(36-38)^. Maintaining Na/K ratios below 2 may represent an intermediary target expected to contribute to reducing blood pressure and CVD risk ^(35, 39)^.

### Salt reduction and health outcomes

A modest reduction in salt intake causes a reduction in blood pressure in both hypertensive and normotensive individuals, with clear public health relevance ^(2, 5)^. There is also emerging evidence that salt reduction and increased use of potassium-based salt substitutes may decrease cardiovascular morbidity and mortality ^(1, 4)^. Addition of salt in meal preparation and consumption on the household and individual level is highly variable, and existing evidence is underpowered to support individual dietary advice on salt reduction to achieve clinical benefits in terms of CVD and mortality ^(40)^. However, pre-processed foods contribute a large proportion of total salt intake and there is a large potential for improving health outcomes through national structural interventions for salt reduction ^(12)^.

### Ongoing action for salt reduction

As part of Norway’s commitment to the global initiative to reduce morbidity and mortality of non-communicable diseases ^(11)^, an action plan for salt reduction was introduced in 2014 ^(41)^, aiming for a reduced population salt intake towards a target of 2.3 g sodium (5.75 g salt) per day ^(42)^. Initiatives include increasing public awareness through communication campaigns, nutrition labelling, monitoring and an intention agreement for salt reduction ^(43)^. The intention agreement on facilitating a healthier diet was signed in 2016 by food industry sectors and the Minister of Health and Care services, thus committing the parties to continue to work towards reducing salt consumption. Between 2014 and 2018 average salt content was successfully reduced in many breads and cereals, while it remained mainly unchanged in most meat and fish products. Overall, 49% of selected food products had a salt content within target levels in 2018, compared with 36% in 2014-15 ^(14)^. Hence, the changes were moderate, and we may also speculate that a shift towards a higher consumption of pre-processed foods might have counteracted the potential benefits of salt reduction of individual foods. Continuous effort by the food industry and catering services has the potential to make a large contribution to reduced population salt intake.

### Strengths and limitations

To our knowledge, this is the first population-based study to examine time trends in estimated salt intake in Norway based on urinary sodium concentrations. The HUNT Study invited all inhabitants in the former Nord-Trøndelag County region and had a reasonably high participation of 54% ^(26)^. The study covered only one region in Norway. However, the population is considered to be fairly representative of Norway except for the lack of large cities and immigrant populations ^(17)^. Data from HUNT used in the international research collaboration *NCD-RisC* show that the population also follows global health trends ^(44)^. The participant sampling, urine collection and biochemical analyses were performed using identical methods in both waves. Another strength is that we had recently validated the INTERSALT equation and found it to perform well in a Norwegian adult population ^(24)^, and the findings in HUNT were comparable to those of that study. Our study had statistical power to detect a reduction in 24-hour salt intake of approximately 0.3 grams or larger. Electrolyte concentrations in a single spot urine sample is an inferior, but more feasible, method for assessing salt intake in populations, as compared to the total sodium excretion measured in 24-hour urine collections. It is thus appropriate for epidemiologic studies aiming at a high participation rate and representativeness. Using spot urine results is reasonable if 24-hour urine samples have been collected from the same population, to achieve the most accurate prediction of 24-hour salt from spot urine ^(20, 23)^. However, studies have suggested a poorer performance of the INTERSALT equation in older adults ^(32)^. Inferences concerning salt consumption in the oldest HUNT4 participants should therefore be drawn with caution. Another limitation of the INTERSALT equation is that it tends to underestimate 24-hour salt at higher intakes and overestimate at lower intakes, with the risk of failing to detect a true decline. However, the unchanged spot urinary sodium concentrations over the period supported our null finding.

In conclusion, we observed no change in estimated 24-hour salt intake in adult men and women in mid-Norway from 2006-08 to 2017-19. Estimated 24-hour salt intakes were lower in women than in men. There was a clear age gradient in women, with the highest mean estimated salt intake in younger adults, but we observed no statistically significant educational gradient in salt intake.

## Data Availability

The data that support the findings of this study are available upon application to the data owner but restrictions apply to the availability of these data, which were used under approval for the purpose of the current research project, and so are not publicly available.

## Acknowledgements

The Trøndelag Health Study (HUNT) is a collaboration between HUNT Research Centre (Faculty of Medicine and Health Sciences, Norwegian University of Science and Technology NTNU), Trøndelag County Council, Central Norway Regional Health Authority, and the Norwegian Institute of Public Health.

