## Supplementary material for "No change in 24-hour salt intake estimated from spot urine in Norwegian adults from 2006 to 2019. The population-based HUNT Study"

**Supplementary Table 1. Characteristics[1] of men and women aged 25-64 years with spot urine samples in the HUNT Study: HUNT3 (2006-08, n=499) and HUNT4 (2017-19, n=500). Median values**

| | Men | | p value[2] | Women | | P value[2] |
|---|---|---|---|---|---|---|
| | HUNT3 | HUNT4 | | HUNT3 | HUNT4 | |
| Age | 45.1 | 45.0 | 0.92 | 44.7 | 45.1 | 0.75 |
| BMI | 27.6 | 27.1 | 0.17 | 25.0 | 25.6 | 0.24 |
| Na (mmol/l) | 121.3 | 114.4 | 0.20 | 86.2 | 82.3 | 0.67 |
| K (mmol/l) | 65.9 | 69.6 | 0.33 | 56.1 | 55.9 | 0.88 |
| Na/K ratio | 1.85 | 1.66 | 0.11 | 1.67 | 1.48 | 0.07 |
| Creatinine | 11.3 | 11.5 | 0.70 | 7.9 | 7.2 | 0.45 |
| Calculated NaCl (g/24h) | 11.10 | 10.83 | 0.14 | 7.58 | 7.59 | 0.92 |

[1] BMI, body mass index. Na, urinary sodium concentration. K, urinary potassium concentration. Na/K ratio, ratio of urinary sodium to potassium concentrations. NaCl, salt.
[2] p-values estimated by quantile regression (Stata function 'qreg')

**Supplementary Table 2. Median spot urinary sodium (Na), potassium (K), sodium to potassium ratio (Na/K ratio), creatinine and estimated 24-hour salt intake by age groups in men and women participating in HUNT4 (n=750).**

| | N | Na (mmol/l) | K (mmol/l) | Na/K ratio | Calculated NaCl (g/24h)[2] | Creatinine (mmol/l) |
|---|---|---|---|---|---|---|
| Men | | | | | | |
| 25-44 years | 125 | 118.5 | 62.4 | 1.99 | 10.6 | 11.5 |
| 45-64 years | 125 | 107.3 | 80.6 | 1.41 | 11.0 | 11.6 |
| 70-79 years | 125 | 102.3 | 68.2 | 1.54 | 11.2 | 10.5 |
| p-value[1] | | 0.10 | 0.24 | **0.003** | **0.017** | 0.35 |
| Women | | | | | | |
| 25-44 years | 95 | 105.8 | 61.8 | 1.65 | 8.0 | 9.1 |
| 45-64 years | 155 | 65.9 | 51.0 | 1.24 | 7.3 | 6.5 |
| 70-79 years[3] | 125 | 85.0 | 61.5 | 1.35 | 5.9 | 6.8 |
| p-value[1] | | **0.027** | 0.74 | **0.043** | **<0.001** | 0.06 |

[1] Age group entered as a continuous variable in quantile regression (Stata function 'qreg')
[2] Estimated using the INTERSALT equation with sex-specific coefficients for the Northern European region (see Supplementary Figure 1)
[3] Three women aged 70-79 years had missing values for BMI and thereby calculated NaCl
P-values <0.05 bolded.

**Supplementary Table 3. Age-standardized median sodium (Na), potassium (K), sodium to potassium ratio (Na/K ratio) and estimated 24-hour salt intake by educational attainment in men and women aged 25-64 years participating in HUNT4.**

| Education[1] | N | Na (mmol/l) | K (mmol/l) | Na/K ratio | Calculated NaCl (g/24h)[3] |
|---|---|---|---|---|---|
| Men | | | | | |
| Primary | 38 | 121.1 | 70.6 | 1.84 | 11.1 |
| Secondary | 110 | 115.0 | 68.4 | 1.82 | 10.8 |
| Tertiary, short (<4 yrs) | 59 | 108.9 | 79.5 | 1.76 | 10.4 |
| Tertiary, long (≥4 yrs) | 42 | 99.2 | 61.8 | 1.50 | 10.7 |
| p-value[2] | | 0.22 | 0.82 | **0.048** | 0.19 |
| Women | | | | | |
| Primary | 39 | 84.6 | 69.3 | 1.54 | 7.8 |
| Secondary | 67 | 100.4 | 53.2 | 1.81 | 8.0 |
| Tertiary, short (<4 yrs) | 72 | 76.1 | 54.9 | 1.37 | 7.5 |
| Tertiary, long (≥4 yrs) | 71 | 81.2 | 55.7 | 1.34 | 7.5 |
| p-value[2] | | 0.38 | 0.43 | **0.020** | **0.007** |

[1] Attained educational level in four categories. Primary education: Completed primary school or 1-2 years of high school/vocational school; secondary education: Completed high school or certificate of apprenticeship; tertiary short: University or other post-secondary education less than 4 years; tertiary long: University/college education of 4 years or more.
[2] Education level entered as a continuous variable in quantile regression (Stata function ‘qreg’)
[3] Estimated using the INTERSALT equation with sex-specific coefficients for the Northern European region (see Supplementary Figure 1)
P-values <0.05 bolded.

**Supplementary Figure 1. The INTERSALT equation with coefficients for the Northern European region for estimating 24-hour sodium excretion (mmol/24h) from spot urine[1,2]**

Men: $25.46 + (0.46*Na_{spot}) – (2.75*Cr_{spot}) – (0.13*K_{spot}) + (4.1*BMI) + (0.26*age) +23.17$

Women: $5.07 + (0.34*Na_{spot}) – (2.16*Cr_{spot}) – (0.09*K_{spot}) + (2.39*BMI) + (2.35*age) – (0.03*age^2) + 15.73$

| | |
|---|---|
| $Na_{spot}$ | Sodium concentration in spot urine, mmol/l |
| $Cr_{spot}$ | Creatinine concentration in spot urine, mmol/l |
| $K_{spot}$ | Potassium concentration in spot urine, mmol/l |
| BMI | Body mass index, kg/m$^2$ |
| Age | Years |

[1] Brown IJ, Dyer AR, Chan Q, Cogswell ME, Ueshima H, Stamler J, et al. Estimating 24-hour urinary sodium excretion from casual urinary sodium concentrations in Western populations: the INTERSALT study. *Am J Epidemiol* 2013;177(11):1180-92

[2] To convert to g Na/24h, results are multiplied by a factor of 0.023, then multiplied by 2.54 to obtain the corresponding g NaCl/24h.
